# Reemergence of Dengue Virus Serotype 3, Brazil, 2023

**DOI:** 10.1101/2023.05.03.23288351

**Authors:** Felipe Gomes Naveca, Gilberto A Santiago, Rodrigo Melo Maito, Cátia Alexandra Ribeiro Meneses, Valdinete Alves do Nascimento, Victor Costa de Souza, Fernanda Oliveira do Nascimento, Dejanane Silva, Matilde Mejía, Luciana Gonçalves, Regina Maria Pinto de Figueiredo, Ana Cecília Ribeiro Cruz, Bruno Tardelli Diniz Nunes, Mayra Marinho Presibella, Nelson Fernando Quallio Marques, Irina Nastassja Riediger, Marcos César Lima de Mendonça, Fernanda de Bruycker-Nogueira, Patricia C Sequeira, Ana Maria Bispo de Filippis, Paola Resende, Tulio Campos, Gabriel Luz Wallau, Tiago Gräf, Edson Delatorre, Edgar Kopp, Andrea Morrison, Jorge L. Muñoz-Jordán, Gonzalo Bello

## Abstract

In 2023, three autochthonous DENV-3 cases were detected in Roraima and one imported case in Paraná, fifteen years after the last DENV-3 outbreak in Brazil. Phylogenetic analyses confirmed all belonging to a new Asian lineage recently introduced in the Americas, raising concerns about future large dengue outbreaks in this region.

## Text

Dengue virus type 3 (DENV-3) comprises five distinct genotypes (I to V). Genotype III (GIII) is the most widespread and was associated with large outbreaks in Asia, Africa, and Americas (1). DENV-3 GIII probably emerged in the Indian subcontinent around the mid-1970s and was first introduced in the Americas in the 1990s. This introduction established an endemic lineage that evolved separately from the Asian lineage for more than 20 years, herein named GIII-American-I, which has been extensively transmitted across the continent in subsequent years (2). The most recent DENV-3 GIII-American-I lineage sequences were reported in Mexico in 2021 (GenBank accession numbers OM417340 and OM417341).

The first autochthonous case of DENV-3 GIII-American-I lineage in Brazil was reported in December 2000 in Rio de Janeiro State (Southeast region) (3,4). This lineage caused a massive dengue outbreak in Rio de Janeiro State in 2002, and subsequent outbreaks were reported in Brazil throughout the 2000s, highlighting its rapid spread (5). Multiple importations of the GIII-American-I lineage from the Lesser Antilles (Caribbean) into Brazil occurred through the 2000s. The Southeast and North Brazilian regions were the most significant dissemination hubs (6,7). In contrast, public data reveals that DENV-3 represented a minor fraction (<1%) of total dengue cases in Brazil since 2010 (**Figure, panel A**), with few cases confirmed by sequencing. Thus, DENV-3 transmission has not been reported for the last few years, suggesting that the DENV-3 GIII-American-I lineage may have become extinct in Brazil.

The present study reports the genetic characterization of four DENV-3 cases detected in Brazil in 2023. Three autochthonous cases were detected by the Roraima State Central Laboratory (LACEN-RR) between 3rd January and 4th March, and one imported case from Suriname was detected in Paraná by the State Central Laboratory (LACEN-PR) on March-2023. Serum samples were sent for sequencing at FIOCRUZ Amazônia, part of the FIOCRUZ Genomics Surveillance Network Consortium of the Brazilian MoH. Four complete DENV-3 genomes were obtained using Illumina technology (VSP), aligned with DENV-3 sequences sampled worldwide, and analyzed for genotyping and spatiotemporal diffusion reconstruction (**Technical Appendix**).

The four complete DENV-3 genomes detected in Brazil in 2023 were classified as GIII according to the Flavivirus Genotyping Tool (https://www.rivm.nl/mpf/typingtool/flavivirus/). Phylogenetic analysis of the DENV-3 GIII dataset revealed that the new GIII sequences detected in Brazil branched together with sequences sampled in Puerto Rico in 2022 and in Florida, USA, in 2022 from both autochthonous and imported cases from Cuba (**Figure 1A**). The new monophyletic clade composed of DENV-3 sequences detected in the Americas between 2022 and 2023, named GIII-American-II, was nested among sequences sampled in Asia belonging to the Asian lineage of GIII over the last decade and outside the GIII-American-I lineage that comprises sequences sampled in the Americas between 1994 and 2021 (**Figure 1A**). Thus, the identified GIII-American-II lineage was not a reemergence of the GIII-American-I lineage previously established in the continent but a new introduction of GIII from Asia. The dissemination of a new DENV-3 lineage in an extensive and populous country like Brazil is concerning because many inhabitants may lack immunity against this serotype. Brazil has not faced recent outbreaks by this serotype; therefore, there is an increased risk of epidemics. Moreover, the endemicity of other DENV serotypes may increase the likelihood of an upsurge of severe cases. The phylogeographic tree estimated for a subset of GIII-American-II sequences and the most closely related GIII Asian sequences (**Technical Appendix**) indicates that this new American lineage was most probably introduced from the Indian subcontinent (posterior state probability [PSP] =1) in 2019 (Bayesian credible interval [BCI] 2018– 2020) and infers that it could have circulated cryptically for ∼3 years before being detected in 2022 (**Figure 1B**). Our phylogeographic analysis points to Cuba as the most probable (PSP = 0.95) introduction point of the GIII-American-II lineage (**Figure 1B**). However, this result is likely biased by the absence of DENV-3 genomes representative of other Caribbean countries. Of note, the estimated onset date of the new GIII-American-II lineage coincides with a severe DENV-3 epidemic described in Jamaica in 2018-2019, the largest dengue outbreak reported on this Caribbean Island in 40 years (8). These observations support the hypothesis that the DENV-3 GIII-American-II lineage might have been introduced from India into the Caribbean region around 2018-2019 and was more recently disseminated from the Caribbean to Brazil, Suriname, and Florida in 2022-2023.

**Figure.**
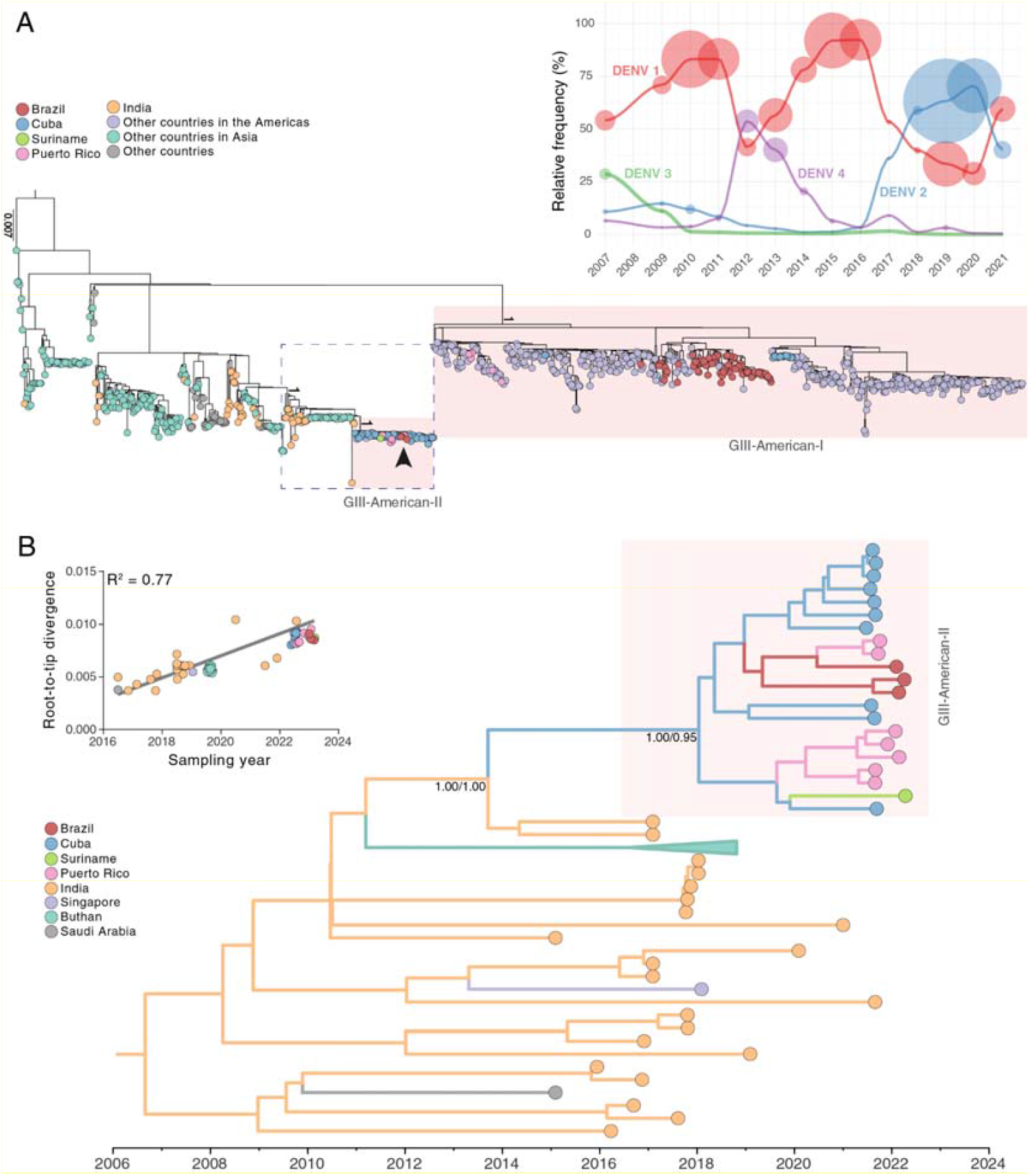
Evolutionary analysis for DENV-3 genotype III, Brazil. A) Rooted tree shows the evolutionary relationships of the DENV-3, genotype III, complete genome sequences of the three autochthonous cases identified in Roraima State and one imported case detected in Paraná State, Brazil (arrow), and 986 publicly available sequences from GenBank. The scale bar indicates nucleotide substitutions per site. Colors represent different sampling locations. Red highlighting shows the two DENV-3 GIII American lineages. The dashed blue rectangle indicates the area enlarged in panel B. Upper-right plot shows the relative frequencies of the four DENV serotypes in Brazil since 2007. The size of the circles is relative to the number of DENV cases attributable to each serotype in each year. B) Time-resolved maximum clade credibility tree showing the blue highlighted area from the larger tree in panel A. Colors indicate geographic sampling location. To improve visualization, the monophyletic clade for Bhutan has been collapsed. Posterior and posterior state probabilities (PP/PSP) values at key nodes show support for branching structure. The inset shows the plot of the root-to-tip genetic distance against sampling time.

In summary, our findings confirm the detection of DENV-3 GIII in the Northern Brazilian region in 2023, associated with a new introduction of this genotype from the Indian sub-continent into the Americas. The > 15-year absence and lack of widespread recent transmission of DENV-3 in Brazil might have rendered the population highly susceptible to infection by this serotype, highlighting the importance of early detection and the need for continuous monitoring of DENV-3 spread in Brazil and other American countries.

## Data Availability

The three new DENV-3 genomes obtained from samples collected in Roraima 2023 are available in GenBank under accession numbers OQ706226-OQ706228. We also deposited another DENV-3 GIII genome from a sample collected in Amazonas 2006 (OQ727062).

## About the Author

Dr. Naveca is a public health researcher at Fiocruz, working in Rio de Janeiro and Amazonas State. His primary research interest lies in diagnostics, molecular epidemiology, and the evolution of emergent and reemergent viruses, mainly arboviruses.

## Author contributions

F.G.N., A.M.B.F., G.A.S., J.L.M.J., and G.B. conceived and designed the study and contributed to data analysis. R.M.M. and C.A.R.M. contributed to diagnostics, patients, and public health surveillance data analysis from Roraima State. M.M.P., N.F.Q.M., and I.N.R. contributed to diagnostics, patient, and public health surveillance data analysis from Paraná State. G.A.S., J.L.M.J., E.K. and A.M. contributed to diagnostics, patient, and sequence data from Puerto Rico and Florida, USA. G.A.S., V.A.N., V.C.S., F.O.N., D.S., M.M., and L.G. contributed with sequencing and data analysis. T.C. contributed to HPC environment configuration and raw sequencing data analysis. FGN, A.M.B.F., and G.B. contributed to laboratory management and obtaining financial support. F.G.N., G.A.S., J.L.M.J., R.M.P.F., A.C.R.C., B.T.D.N., M.C.L.M., F.B-N, P.C.S., P.C.R., G.L.W., E.D., T.G., and G.B. contributed to epidemiological and phylogenetic data analysis. F.G.N, V.A.N., G.L.W., T.G., E.D. G.B. wrote the first draft, and all authors contributed and approved the final manuscript.

## Technical Appendix

### Materials and Methods

#### Ethics statement, sample collection, and arboviral detection as a surveillance routine in Central State Laboratories

In Roraima State and Paraná State most arboviral suspected cases are investigated for Zika, Dengue, and Chikungunya (ZDC) by the State’s Central Laboratory (LACEN-RR in Roraima and LACEN-PR in Paraná) as a routine. Between 1st January and 27th March, 319 suspected acute arboviral samples were sent to the Roraima State Central Laboratory (LACEN-RR) for molecular diagnosis. At LACEN-RR, these samples underwent a nucleic acid extraction with commercial kits, followed by ZDC detection using the reverse transcriptase real-time PCR kit developed by Instituto de Biologia Molecular do Paraná (IBMP - Kit Biomol ZDC - https://www.ibmp.org.br), which not only detects DENV but also identifies its serotype. Only 10 cases were confirmed as dengue (DENV-2 = 4, DENV-3 = 4) or chikungunya (n = 2) by real-time reverse transcription PCR. Clinical and epidemiological data, plus IgM detection, confirmed 19 other cases (DENV = 9, Chikungunya = 10). The socio-demographic and laboratory characteristics of the three DENV-3 cases are summarized in supplemental Table 1. The case from Paraná described in the present manuscript is considered as an imported case from Suriname since the patient arrived in Brazil with dengue symptoms three days before the sample collection. This study was approved by the Ethics Committee of Instituto Oswaldo Cruz, which waived signed informed consent (CAAE: 90249218.6.1001.5248).

#### Epidemiological data collection and visualization

Epidemiological data on dengue fever cases in Brazil were downloaded from the DATASUS database using the R package microdatasus (1). Specifically, data on the number of confirmed DENV cases and their corresponding serotypes were obtained. However, serotype information was only available from 2007 onwards. The data were extracted at the national level and stratified by year. Plots were produced using the ggplot2 package on R software version 4.1.2.

#### Nucleotide sequencing library preparation

As requested by local health authorities, DENV-3 positive serum samples were sent to Instituto Leônidas and Maria Deane (ILMD - Fiocruz Amazônia) for nucleotide sequencing and genomics characterization. At ILMD, samples were submitted to viral total nucleic acid extraction with a commercial kit (Promega Maxwell), according to the manufacturer’s instructions. Thus, the total nucleic acid was used for the sequencing library with Illumina’s Viral Surveillance Panel (VSP w ILMN RNA Prep w Enrich). The VSP manual (# 1000000124435 v03, April 2021) describes the preparation of viral genomic libraries through a hybrid capture method using biotinylated probes and enrichment for the complete genome sequencing of 66 human pathogens, including DENV-3, allowing the detection of mutations and new lineages, without bias.

#### Whole-genome sequencing and consensus genomes

The VSP libraries were then submitted to nucleotide sequencing on a MiSeq instrument using V3 cartridges on 2□×□150 cycles paired-end run (Illumina). The FastQ files generated at Illumina’s cloud (https://basespace.illumina.com) were submitted to further quality check and duplicate removal steps using a customized workflow employing BBDuk (v38.84). The consensus genomes were assembled into contigs using BBMap and the GenBank DENV-3 reference genome (NC_001475) as the template. We used the BBTools (BBDuk and BBMap v38.84 - https://sourceforge.net/projects/bbmap/) versions embedded in Geneious Prime 2023.0.4 software (https://www.geneious.com). Finally, all contigs were carefully visually inspected before further analysis. The obtained consensus genomes ranged in length from 10,511 to 10,697 nucleotides, or 98.2 to 99.9% of the DENV-3 RefSeq, with no ambiguous or unidentified sites (“N”).

#### DENV-3 genotype identification

The DENV-3 consensus genomes obtained in this study were initially submitted to genotype identification using the Flavivirus Genotyping Tool Version 0.1 (https://www.rivm.nl/mpf/typingtool/flavivirus/). This analysis showed that the three genomes belong to the DENV-3 genotype III, with high bootstrap support (100.0).

#### DENV-3 dataset and global phylogenomics reconstruction

We downloaded all DENV-3 genomes (genome length > 10K) available on GenBank (2023-03-25) using the NCBI Virus portal (https://www.ncbi.nlm.nih.gov/labs/virus/vssi/#/). Subsequently, we confirmed both the serotype (DENV-3) and the genotype (GIII) using the same Flavivirus genotyping web tool previously mentioned, followed by removing sequences named as modified nucleic acids or linked to patents. Thus, this first dataset encompasses 986 DENV-3 (GIII) genomes available at GenBank, which were aligned with the four genomes obtained in this study using MAFFT v7.490 under the automatic selection of the appropriate strategy according to data size (2).

The aligned dataset was used to reconstruct phylogenomic relationships by maximum likelihood (ML) using IQ-TREE multicore version 2.0.3 (3). All sequences passed the IQ-TREE composition test, and ModelFinder (4) chose the GTR+F+R4 evolutionary model according to Bayesian Information Criterion. The tree branches’ support was evaluated with Shimodaira–Hasegawa approximate likelihood-ratio test (SH-aLRT) (5) and Ultrafast bootstrap (UFBoot) (6), with 2,000 replicates.

#### Spatiotemporal and viral diffusion reconstruction of DENV-3

To reconstruct the geographic and temporal origin of DENV-3 cases in Brazil in 2023, we employed a Bayesian phylogeographic inference approach implemented in BEAST v1.10.4 (7). To reduce computation time in the phylogeographic reconstructions, we selected the sub-set of most closely related Asian sequences to the newly generated DENV-3 genomes, as observed in the ML analysis. Moreover, we generate a “non-redundant” sub-set of 10 DENV-3 sequences representative of the viral diversity in imported cases from Cuba detected in Florida. To achieve this aim, sequences imported from Cuba were grouped by similarity (genetic similarity > 0.985) with the CD-HIT program (8), and only one sequence per cluster (the oldest one) was selected. Finally, we exclude potential recombinant genomes detected by at least five out of seven different algorithms (RDP, GENECONV, Chimaera, MaxChi, BootScan, SiScan, and 3seq) running in RDP5.34 (9) and temporal outlier sequences that deviated more than 1.5 interquartile ranges in the regression analysis of the root-to-tip divergence against the tip sampling time of the ML phylogenetic tree performed using TempEst v.1.5.3 (10). Time-scaled trees were inferred with a relaxed molecular clock model (11), outperforming the strict clock model in marginal likelihood estimation and a Bayesian Skyline coalescent model. The ancestral node states were reconstructed with a continuous-time Markov chain (CTMC) prior (12) and discrete spatial diffusion with an asymmetric substitution model to infer the migration events (13). Bayesian stochastic search variable selection (BSSVS) was performed to identify the significant migration routes. Two Markov Chain Monte Carlo (MCMC) chains were run for 100 million generations and combined after removing 10% burn-in. Convergence was assessed by calculating the Effective Sample Size (ESS) for all parameters using Tracer v1.7.1 (14). The time-scaled phylogeny was visualized using FigTree v1.4.4 (http://tree.bio.ed.ac.uk/software/figtree).

## Data availability

The four new DENV-3 genomes obtained from samples collected in Brazil in 2023 are available in GenBank under accession numbers OQ706226-OQ706228 (Roraima State 2023) and OQ868517 (Paraná State, imported from Suriname). We also deposited another DENV-3 GIII genome obtained from a sample collected in Amazonas 2006 (OQ727062).

## Acknowledgments

We acknowledge all data contributors, i.e., authors and their originating laboratories responsible for obtaining and submitting timely Dengue virus genomes, including metadata, via GenBank or other public databases. We appreciate the support of FIOCRUZ Genomics Surveillance Network members, as well as the Roraima State and Paraná State surveillance teams for the partnership in viral surveillance in Brazil. We would also like to thank Claudia Nunes Duarte dos Santos for critically reading the manuscript. Funding support: FAPEAM (Universal/AM call 2019; Rede Genômica de Vigilância em Saúde - REGESAM); Inova Fiocruz/Fundação Oswaldo Cruz (Inova Amazônia); Departamento de Ciência e Tecnologia (DECIT) of the Brazilian MoH; CNPq through their productivity research fellowships (306146/2017–7, 303902/2019–1 and 304883/2020-4, respectively): F.G.N., G.L.W., and G.B., respectively. FAPERJ (Grant number E-26/202.896/2018).

---

**Supplemental Table 1.** Socio-demographic and laboratory characteristics of the DENV-3 cases, Brazil, 2023.

| **GenBank Accession** | **Brazilian State** | **Municipality** | **Collection** | **Age range** | **Gender** | **Symptoms** | **Ct** | **Genome length** |
| --- | --- | --- | --- | --- | --- | --- | --- | --- |
| OQ706226 | Roraima | Cantá | March-2023 | 21-25 | Female | F, M | 31 | 10,697 |
| OQ706227 | Roraima | Boa Vista | January-2023 | 6-10 | Male | F, L, P, PE, and T | 28 | 10,511 |
| OQ706228 | Roraima | Boa Vista | January-2023 | 51-55 | Female | F, BP, M, R, and V | 29 | 10,553 |
| OQ868517 | Paraná | Curitiba* | March-2023 | 31-35 | Male | F, Fa, H, M, N, R, and V | 27 | 10,673 |

Symptoms: BP = back pain, F= Fever, Fa = fatigue, H = headache, L = leucopenia, M = myalgia, N= nausea, P = polyarthralgia, PE = periarticular edema, R = rash, T = thrombocytopenia, V = vomiting. Ct = Cycle threshold. \* Although the sample was collected in Curitiba, Paraná, Brazil, the patient arrived in Brazil on March 2023 with dengue symptoms, thus it is considered by local authorities as an imported case from Suriname.

